# Real-world evidence of triplet therapy in metastatic hormone-sensitive prostate cancer - an Austrian multicenter study

**DOI:** 10.1101/2023.10.13.23297000

**Authors:** Mona Kafka, Giulia Giannini, Nastasiia Artamonova, Hannes Neuwirt, Heidemarie Ofner, Gero Kramer, Thomas Bauernhofer, Ferdinand Luger, Thomas Höfner, Wolfgang Loidl, Hubert Griessner, Lukas Lusuardi, Antonia Bergmaier, Andreas Berger, Thomas Winder, Sarah Weiss, Severin Bauinger, Steffen Krause, Martin Drerup, Elmar Heinrich, Magdalena Schneider, Stephan Madersbacher, Sonia Vallet, Franz Stoiber, Sarah Laimer, Stephan Hruby, Gert Schachtner, Udo Nagele, Sebastian Lenart, Anton Ponholzer, Jacob Pfuner, Clemens Wiesinger, Christoph Kamhuber, Ecan Müldür, Wolfgang Horninger, Isabel Heidegger

## Abstract

**Introduction:** Two randomized trials demonstrated a survival benefit of triplet therapy (androgen deprivation therapy [ADT]) plus androgen receptor pathway inhibitor [ARPI] plus docetaxel) over doublet therapy (ADT plus docetaxel) changing treatment strategies in metastatic hormone-sensitive prostate cancer (mHSPC).

**Patients and methods:** We conducted the first real-world analysis including 97 mHSPC patients from sixteen Austrian medical centers. 79.4% of patients received abiraterone, 17.5% darolutamide, 2.1% apalutamide and 1% enzalutamide. Baseline characteristics and clinical parameters during triplet therapy were documented. Mann-Whitney-U-Test for continuous or X²-test for categorical variables was used. Variables on progression were tested using logistic regression analysis and tabulated as hazard ratios (HR), 95% confidence interval (CI).

**Results:** 83.5% of patients with synchronous and 16.5% with metachronous disease were included, with 83.5% high-volume disease diagnosed by conventional imaging (48.9%) or PSMA PET-CT (51.1%). While docetaxel and ARPI were administered consistent with pivotal trials, prednisolone, prophylactic gCSF and osteoprotective agents were not applied guideline conform in 32.5%, 37% and 24.3% of patients, respectively. Importantly, a non-simultaneous onset of chemotherapy and ARPI, performed in 44.8% of patients, was significantly associated with worse treatment response (p=0.015, HR 0.245). Starting ARPI before chemotherapy was associated with significant higher probability for progression (p=0.023, HR 15.781) than vice versa. Strikingly, 15.6% (abiraterone) and 25.5% (darolutamide) low-volume patients as well as 14.4% (abiraterone) and 17.6% (darolutamide) metachronous patients received triplet therapy. Adverse events (AE) occurred in 61.9% with grade 3-5 in 15% of patient without age-related differences. All patients achieved a PSA decline of 99% and imaging response was confirmed in 88% of abiraterone and 75% of darolutamide patients.

**Conclusions:** Triplet therapy arrived in clinical practice primarily for synchronous high-volume mHSPC. Regardless of selected therapy regimen, treatment is highly effective and tolerable. Preferably therapy should start simultaneously, if not possible chemotherapy should be started first.

**Take Home Massages:** Triplet therapy consisting of ADT plus ARPI (abiraterone or darolutamide) plus docetaxel is an effective and mostly well tolerable treatment option for mHSPC patients also in the real-world setting especially for synchronous, high-volume patients.

---

However, in the real-world setting, triplet therapy is not started simultaneously in about 50% of cases leading to decreased treatment response.

## 1. Introduction

Despite many therapeutic advances in recent years, the majority of hormone-sensitive prostate cancer (mHSPC) patients become castration resistant (mCRPC) associated with high morbidity and mortality. Thus, the primary goal of mHSPC treatment is to prolong survival through long-term tumor suppression by targeting the key drivers of carcinogenesis with the androgen receptor (AR) pathway as a crucial factor (1,2). While classical androgen deprivation therapy (ADT) decreases the amount of androgen, androgen receptor pathway inhibitors (ARPIs) act by inhibiting androgen synthesis (abiraterone) or by competitively binding to the AR (apalutamide, darolutamide, enzalutamide) (3–7). Presently, guidelines recommend a dual combination consisting of ADT plus ARPI or ADT plus docetaxel chemotherapy depending on patient characteristics and prognostic factors including occurrence of metastases (synchronous vs. metachronous) and metastatic load (high vs. low-volume according to CHAARTED criteria) (8,9). While patients with low-volume metachronous mHSPC have the best prognosis (5-year overall survival (OS) 70–75%), high-volume synchronous mHSPC is associated with the worst prognosis (5-year OS 20–30%) claiming for intensified personalized treatment (10).

Two recent randomized controlled trials (RCTs) demonstrated an OS benefit if mHSPC patients were treated with triplet therapy consisting of ADT plus ARPI plus docetaxel, introducing the concept of treatment intensification by combining hormonal therapy with cytotoxic chemotherapy in prostate cancer (PCa) (11,12). Briefly, PEACE-1 compared ADT plus docetaxel plus abiraterone to ADT plus docetaxel in 710 synchronous mHSPC patients demonstrating that triplet therapy increased OS by almost 25%, especially in high-volume disease with a median OS of 5.1 years vs. 3.5 years (11). In the ARASENS trial, mHSPC patients received ADT plus docetaxel plus darolutamide or ADT plus docetaxel plus placebo. This regime resulted in a reduction of the risk of death by approximately 32%. Additionally, ARASENS included metachronous patients (13.2% treatment, 12.5% control) demonstrating also a survival benefit in those patients (12). Although PEACE-1 was the first trial demonstrating a benefit of triplet therapy, in January 2023 darolutamide (Nubeqa®) was approved for mHSPC in combination with ADT and chemotherapy based on the ARASENS trial. Hence, current 2023 EAU guidelines already recommend both drugs in triplet therapy combinations for patients eligible for docetaxel who are willing to accept the increased risk of side effects (8).

Currently, no real-world data exist delineating clinical performance of triplet therapy outside registration trials. Thus, we conducted this first real-world analysis assessing the implementation of triplet therapy in clinical practice as well as its efficacy and tolerability.

## 2. Patients and Methods

### 2.1 Patient cohort

After obtaining a positive vote of the local ethical committees a retrospective multicenter study was performed. A total of 97 men diagnosed with synchronous or metachronous mHSPC derived from 16 Austrian medical centers were included. All patients had an Eastern Cooperative Oncology Group (ECOG) performance status of 0 or 1 and were eligible to receive a triple therapy consisting of ADT plus docetaxel plus ARPI (abiraterone or darolutamide). One patient received enzalutamide, based on findings of the ENZAMET trial (13).

### 2.2 Statistical analysis

Statistical analysis was performed using SPSS, version 26.0 (IBM Corp., Armonk, NY, USA). Baseline characteristics were used to describe the population. Median, minimum and maximum were used for continuous variables, proportions are depicted as numbers and percentages. To calculate differences between groups Mann-Whitney-U-Test for continuous or the X²-test for categorical variables was used. The influence of different variables on progression was tested using uni- and multivariate logistic regression analysis and tabulated as hazard ratios (HR), 95% confidence interval (CI). All statistical tests were two-sided at a significance level of p<0.05.

## 3. Results

### 3.1 Patient characteristics

A total of 97 patients with a median age of 68 years from sixteen Austrian centers treated by urologists (n=57, 55.3%), oncologists (n=23, 22.3%) or both (n=17, 22.4%) were included in this retrospective multicenter observational study. 5/16 centers were classified as academic institutions, 8/16 hospitals were high volume case centers.

83.5% of our patients harbored a synchronous and 16.5% a metachronous disease. 83.5% of men were classified as high-volume according to the CHAARTED criteria diagnosed either by conventional imaging using contrast-enhanced computed tomography (CT) and bone scan (48.9%) or PSMA PET-CT (51.1%). In 51.5% a Gonadotropin-Releasing-Hormone (GnRH) agonist was applied as ADT and in 48.5% an antagonist. Of importance, 79.4% of patients were treated according to PEACE-1 using abiraterone, while in 17.5% darolutamide was administered in line with the ARASENS protocol. One patient was treated by enzalutamide based on a subgroup analysis of ENZAMET and in two patients apalutamide was used.

Of importance 13/16 of metachronous mHSPC patients were treated with abiraterone contrary to PEACE-1 protocol where only synchronous patients were included. All patients received the full dose of ARPI implicating 1000 mg/d abiraterone, 600 mg/d darolutamide or 240 mg/d enzalutamide. In 94.8% docetaxel was applied in line with RCTs, (75 mg/m^2^ of body surface area (BSA) every 21 days for 6 cycles) and initiated within three months after the start of ADT in 95.4% of patients (**Table 1**).

**Table 1:** Patient characteristics at primary diagnosis. ARPI= Androgen receptor pathway inhibitor, PSA= prostate specific antigen, ISUP= International Society of Urological Pathology, BSA= body surface area, GnRH= Gonadotropin-Releasing-Hormone, BRCA= Breast Cancer gene; *primary diagnosis by metastatic biopsy, no ISUP available

| Parameter | N (%) | Median | Mean | Min-Max |
| --- | --- | --- | --- | --- |
| **Age at diagnosis (years)** |  | 68 | 66 | 49-94 |
| **PSA at primary diagnosis (ng/ml)** |  | 52.5 | 686.34 | 1.8-35278 |
| **ISUP classification** |  |  |  |  |
| - ISUP 1 | 1 (1%) |  |  |  |
| - ISUP 2 | 6 (6.2%) |  |  |  |
| - ISUP 3 | 8 (8.2 %) |  |  |  |
| - ISUP 4 | 28 (28.9%) |  |  |  |
| - ISUP 5 | 53 (54.6%) |  |  |  |
| - Not applicable* | 1 (1%) |  |  |  |
| **Volume of disease (CHAARTED)** |  |  |  |  |
| - Low | 16 (16.5%) |  |  |  |
| - High | 81 (83.5%) |  |  |  |
| **Type of metastatic disease** |  |  |  |  |
| - Synchronous | 81 (83.5%) |  |  |  |
| - Metachronous | 16 (16.5%) |  |  |  |
| **Location of metastasis** |  |  |  |  |
| - Bone only | 20 (20.6%) |  |  |  |
| - Lymph nodes only | 5 (5.2%) |  |  |  |
| - Visceral only | 2 (2.1%) |  |  |  |
| - Multiple sites | 70 (72.2%) |  |  |  |
| **Imaging type at primary diagnosis** |  |  |  |  |
| - CT + Bone scan | 38 (39.5%) |  |  |  |
| - PSMA PET-CT | 38 (39.5%) |  |  |  |
| - CT only | 6 (6.3%) |  |  |  |
| - CT + Bone scan + PSMA PET | 8 (8.3%) |  |  |  |
| - Choline PET or FDG PET | 7 (7.2%) |  |  |  |
| **Imaging used for classification of volume** |  |  |  |  |
| - CT + Bone scan | 45 (48.9%) |  |  |  |
| - PSMA PET-CT | 47 (51.1%) |  |  |  |
| **Prostate cancer familiarity** |  |  |  |  |
| - Yes | 8 (9.9%) |  |  |  |
| - No | 73 (90.1%) |  |  |  |
| **BRCA 1/2 mutation** |  |  |  |  |
| - Yes | 4 (4.1%) |  |  |  |
| - No | 27 (27.8%) |  |  |  |
| - Not tested | 66 (68%) |  |  |  |
| **ADT** |  |  |  |  |
| - GnRH Agonist | 50 (51.5%) |  |  |  |
| - GnRH Antagonist | 47 (48.5%) |  |  |  |
| **ARPI** |  |  |  |  |
| - Abiraterone | 77 (79.4%) |  |  |  |
| - Darolutamide | 17 (17.5%) |  |  |  |
| - Enzalutamide | 1 (1%) |  |  |  |
| - Apalutamide | 2 (2.1%) |  |  |  |
| **Start of triplet within 3 months after ADT start** |  |  |  |  |
| - Yes | 93 (95.9%) |  |  |  |
| - No | 4 (4.1%) |  |  |  |
| **Docetaxel** |  |  |  |  |
| - 75 mg/m<sup>2</sup>/BSA/6 cycles 3 weekly | 92 (94.8%) |  |  |  |
| - 50 mg/m<sup>2</sup>/BSA/8 cycles 2 weekly | 5 (5.2%) |  |  |  |

### 3.2 Mode of application in the real-world setting

In 44.8% of patients chemotherapy and ARPI were started concomitantly. In 55.2% treatment was started with a time delay, among them in 79.2% (n=42/53) ARPI was started first. Specifying the time delay, in 45.6% combination therapy was started within one week, however in 15.6% the onset delayed for more than 28 days (**Table 2**). Assessing concomitant supportive medication, we found that only 50% of patients sustained prophylactic granulocyte colony-stimulating factor (gCSF) 24 hours after chemotherapy. Despite lack of recommendation in all guidelines, 27.9% of bone metastatic patients received osteoprotection by denosumab 120 mg/sc. monthly. Assessing the prednisolone dose in patients treated according to PEACE-1, 62.3% received guideline conform 10 mg prednisone/day. In addition, 7.8% (n=7/77) of abiraterone patients had diabetes mellitus as comorbidity. Since abiraterone has become generic in October 2022, we evaluated the use of generics and found that 50% of patients who started a PEACE-1 based triplet therapy were treated with a generic (**Table 2**).

**Table 2:** Assessment of triplet therapy administration. ARPI= Androgen receptor pathway inhibitor, gCSF= granulocyte colony-stimulating factor

| Parameter | N (%) |
| --- | --- |
| **Interval Chemotherapy - ARPI** |  |
| Simultaneously | 43 (44.8%) |
| Delayed | 53 (55.2%) |
| - Chemotherapy - ARPI | 11 (20.8%) |
| - ARPI - Chemotherapy | 42 (79.2%) |
| Time delay (days) |  |
| - 1-7 days | 41 (45.6%) |
| - 8-14 days | 15 (16.7%) |
| - 15-21 days | 7 (7.8%) |
| - 22-28 days | 13 (14.4%) |
| - > 28 days | 14 (15.6%) |
| **Disease volume - Abiraterone** |  |
| - Low | 12 (15.6%) |
| - High | 65 (84.4%) |
| **Disease volume - Darolutamide** |  |
| - Low | 4 (23.5%) |
| - High | 13 (76.5%) |
| **Metastatic stage - Abiraterone** |  |
| - Synchronous | 66 (85.7%) |
| - Metachronous | 11 (14.3%) |
| **Metastatic stage - Darolutamide** |  |
| - Synchronous | 14 (82.4%) |
| - Metachronous | 3 (17.6%) |
| **gCSF during chemotherapy** |  |
| - Yes | 51 (52.3%) |
| - No | 37 (38.1%) |
| - Unknown | 9 (9.3%) |
| **Prednisolone dose - Abiraterone** |  |
| - 5mg | 23 (29.9%) |
| - 10 mg | 48 (62.3%) |
| - No prednisolone | 2 (2.6%) |
| - Unknown | 4 (5.2%) |
| **Osteoprotection - osseous disease** |  |
| - Yes | 27 (27.9%) |
| - No | 63 (64.9 %) |
| - Unknown | 7 (7.2%) |

While PEACE-1 data were first presented at the ASCO meeting 2021, data from ARASENS were presented seven months later at the ASCO GU meeting 2022. Nonetheless darolutamide has already been both FDA (08/2022) and EMA (01/2023) approved. Interestingly, we observed that since the EMA approval 71% in our cohort were treated with darolutamide and 29% with abiraterone respectively, suggesting that darolutamide is the preferred ARPI in triplet therapy regimes.

As in contrast to abiraterone, darolutamide has been shown its efficacy in both low-volume and metachronous mHSPC patients, we tabulated those characteristics in our collective. As assumed, most patients belong to the high-volume group, however 15.6% and 23.5% of low-volume patients were treated with abiraterone or darolutamide. Analyzing our patients with metachronous disease, we depicted treatment with abiraterone and darolutamide in 14.4% and 17.6%, respectively.

Calculating differences in ARPI treatment based on clinical variables including age, PSA, ISUP, disease volume as well as metastasis, we did not find significant differences among treatment groups (**Supplementary Table 1**).

### 3.3 Safety

Incidence of any adverse events (AE) across patients treated with ARPIs was 61.9% including 15% patients with grade 3-5 AE (n=15/97), with no age-related differences stratifying patients according to groups (≤ 65 vs. >65 years). 14.3% of patients did not receive full 6 cycles of chemotherapy due to side effects (n=14/97). The most frequently reported AE were fatigue, polyneuropathy and dermatological disorders including skin rash, hand and foot syndrome, pruritus, hypersensitivity reaction, stomatitis, alopecia and nail pigmentation. Febrile neutropenia as a special AE occurred in 6/97 patients (5.8%), of which only 50% of patients received prophylactic gCSF application. Evaluating the need of a local treatment during triplet therapy, 23.7% had a surgical treatment of the urinary tract due to local complications (**Table 3**).

**Table 3:** Side effects during triplet therapy across all ARPIs: AE=adverse event 1 skin rash, hand and foot syndrome, pruritus, hypersensitivity reaction, stomatitis, alopecia, nail pigmentation, hypersensitivity reaction ^2^ bronchitis, candida endophtalmitis, pneumonitis, pneumonia, infection not classified ^3^ cephalea, taste disorder, sleep disorders, anxiety, dizziness, restless legs ^4^ nausea, diarrhea, increased AST/ALT level

| Parameter | N (%) |
| --- | --- |
| **Any AE** |  |
| - Yes | 60 (61.9%) |
| - No | 28 (28.9%) |
| - Unknown | 9 (9.3%) |
| **Age related AE** |  |
| ≤ 65 years |  |
| - Yes | 25 (61%) |
| - No | 14 (34.1%) |
| - Unknown | 2 (4.9%) |
| >65 years |  |
| - Yes | 34 (62.9) |
| - No | 14 (25.9%) |
| - Unknown | 6 (11.1%) |
| **AE Grading** |  |
| - I | 24 (40%) |
| - II | 16 (26.7%) |
| - III | 12 (20%) |
| - IV | 6 (10%) |
| - V (death) | 2 (3.3%) |
| AE leading to discontinuation of triplet therapy | 14 (14.3%) |
| **Specification of side effects** |  |
| - Fatigue | 15 (18.5%) |
| - Dermatologic<sup>1</sup> | 13 (16%) |
| - Infection<sup>2</sup> | 5 (6.2%) |
| - Sepsis | 2 (2.5%) |
| - Multi-organ failure | 2 (2.5%) |
| - Neutropenia | 4 (4.9%) |
| - Febrile Neutropenia | 6 (7.4%) |
| - Hypokalemia | 1 (1.2%) |
| - Polyneuropathy | 14 (17.3%) |
| - Nervous system<sup>2</sup> | 9 (11.1%) |
| - Gastrointestinal | 4 (4.9%) |
| - Peripheral edema | 3 (3.7%) |
| - Cardiac decompensation | 1 (1.2%) |
| - Adrenal insufficiency | 1 (1.2%) |
| - Stroke | 1 (1.2%) |
| **Local treatment** |  |
| - None | 63 (64.9%) |
| - Urinary catheter | 3 (3.1%) |
| - Nephrostomy | 6 (6.2%) |
| - Transurethral resection of the prostate | 2 (2.1%) |
| - Cytoreductive radical prostatectomy | 2 (2.1%) |
| - Radiation prostate | 9 (9.3%) |
| - Other | 1 (1%) |
|  | 11 (11.3%) |
| - Unknown |  |

Next, we categorized AEs according to ARPI to elucidate if one is better tolerable. Summarizing the incidences of the most common AEs, many of them are known toxic effects related to docetaxel, AEs were similar in both groups with serious events occurring in 62.3% of patients in the abiraterone and 58.8% in the darolutamide group. Of importance, two patients died due to multi-organ failure, one patient in each triple therapy group (abiraterone/darolutamide). Stratifying side effects during triplet therapy according to selected ARPI, no significant differences in the occurrence of AE as well as in AE severity was observed. Therefore, side effects of both ARPIs seem quite comparable in the triple therapy setting (**Table 4**).

**Table 4:**
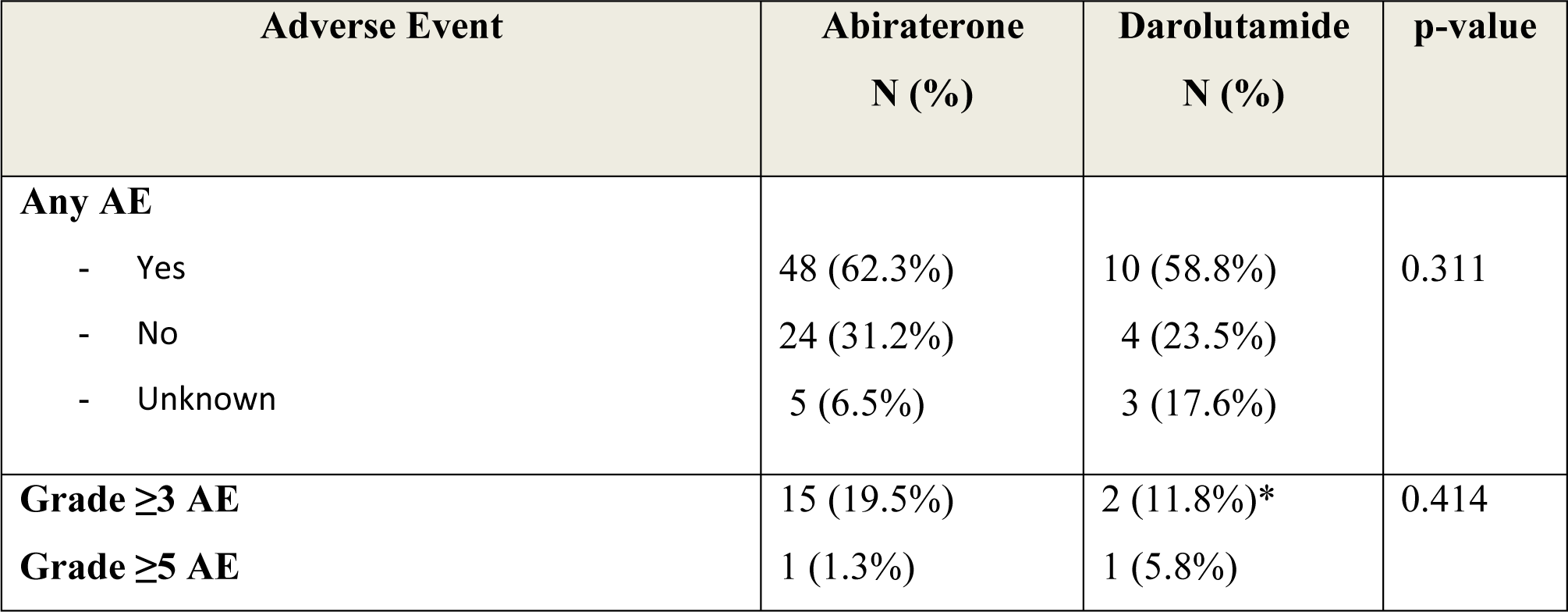
Side effects during triplet therapy according to ARPI; abiraterone (n=77) or darolutamide (n=17) *Neutropenia

| Adverse Event | Abiraterone<br>N (%) | Darolutamide<br>N (%) | p-value |
| --- | --- | --- | --- |
| **Any AE** |  |  |  |
| - Yes | 48 (62.3%) | 10 (58.8%) | 0.311 |
| - No | 24 (31.2%) | 4 (23.5%) |  |
| - Unknown | 5 (6.5%) | 3 (17.6%) |  |
| **Grade $\geq 3$ AE** | 15 (19.5%) | 2 (11.8%)* | 0.414 |
| **Grade $\geq 5$ AE** | 1 (1.3%) | 1 (5.8%) |  |

### 3.4 Efficacy of triplet therapy and clinical predictor for treatment response

Next, we evaluated treatment efficacy of triplet therapy to get a first impression on its performance in the real-life setting. In our cohort all patients had a PSA decline of 99%, and also a response on imaging was confirmed in 81% of patients in the abiraterone and in 75% of darolutamide patients, respectively. Long term follow-up data calculated for abiraterone patients revealed a time to progress and time to castration resistance of 8.5 months and a time to death of 14.7 months (**Table 5**).

**Table 5:** Evaluation of treatment efficacy: standard deviation (SD), *\*abiraterone* patients only

| Parameter | Abiraterone | Darolutamide | p-value |
| --- | --- | --- | --- |
| **PSA decrease % (SD)** | 99.94% (2.97) | 99.86% (0.3) | 0.155 |
| **PSA nadir (ng/ml) Median (SD)** | 0.04 (3.86) | 0.13 (0.14) | 0.549 |
| **Imaging response N (%)** |  |  |  |
| - Progressive disease | 1 (1.7%) | 0 | 0.561 |
| - Stable disease | 10 (17.2%) | 2 (25%) |  |
| - Response | 41 (70.7%) | 4 (50%) |  |
| - Mixed response | 6 (10.3%) | 2 (25%) |  |
| **Response N (%)** |  |  |  |
| - Stable disease/ Response | 51 (88%) | 6 (75%) | 0.06 |
| - Progressive disease/Mixed response | 7 (12%) | 2 (25%) |  |
| **Time to progression months (SD)\*** | 8.5 (4.65) |  |  |
| **Time to castration resistance** *months (SD)\** | 8.5 (3.54) |  |  |
| **Time to 2°-line therapy** *months (SD)\** | 8 (3.51) |  |  |
| **Time to death** *months (SD)\** | 14.7 (11.3) |  |  |

Evaluating clinical predictors for treatment response we confirmed that synchronous disease is associated with worse survival than metachronous disease (p<0.001). In addition, we showed that diabetic patients (p=0.003) and patients without full-dose chemotherapy (p=0.01) had worse survival rates. Furthermore, we assessed tumor- and patient-based characteristics associated with treatment response to triplet therapy stratified according to the administered ARPI (**Table 6**).

**Table 6:**
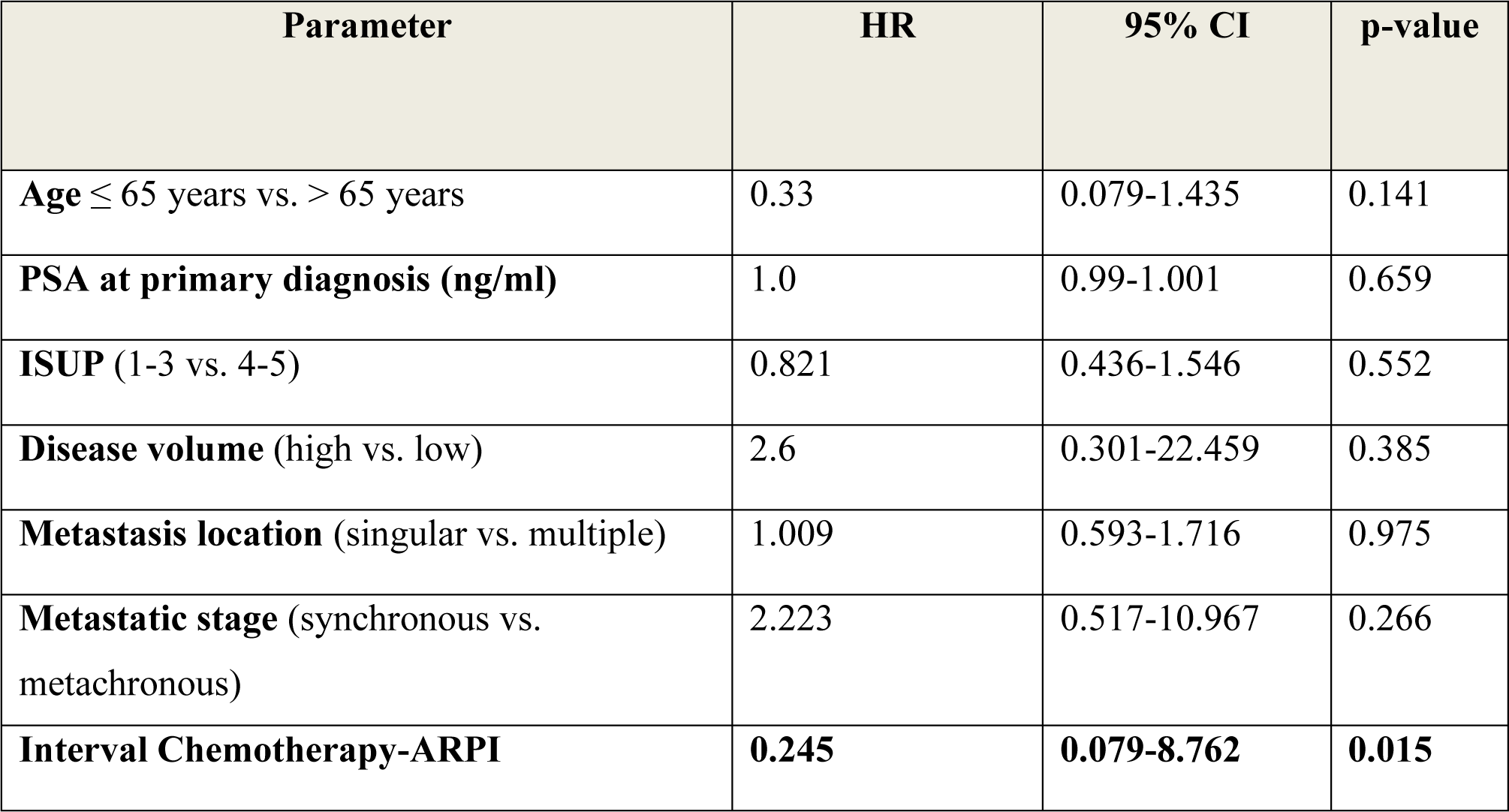
Clinical parameters stratified according to the abiraterone vs. darolutamide group predicting image-based therapy response (Progressive disease and mixed response vs. stable disease and response): Hazard ratio (HR), Confidence interval (CI)

Of utmost importance, a time delay in treatment start of chemotherapy and ARPI was significantly associated with worse treatment response (p=0.015, HR 0.245) (**Table 6**). Assessing the sequence, starting ARPI before chemotherapy was associated with a significant higher probability for progression after chemotherapy (p=0.023, HR 15.781), while chemotherapy followed by ARPI did not influence treatment response (p=0.256, HR 3.703). Summarizing, our data clearly show that I) triplet therapy should be started simultaneously, and II) if a time delay is not evitable, chemotherapy should be started first.

## 4. Discussion

We present the first analysis of the clinical efficacy and tolerability of triplet therapy in mHSPC patients in the real-world setting. Summarizing our findings we demonstrate that I) mostly synchronous metastatic young patients with high tumor load are treated with triplet therapy; II) in half of the cases therapy is not started simultaneously associated with significant higher risk of progression; III) triplet therapy is highly effective, PSA based response rates of 99% and imaging response in 81% of abiraterone and 75% of darolutamide patients, respectively; IV) Grad ≥III side effects occurred in 1/3 of patients including two Grad V events with no differences among treatment groups.

Our patient collective comprised 83.5 % of patients with high-volume disease suggesting that triplet combinations are offered significantly preferably to patients with high metastatic load. Worth mentioning, 15.6 % of patients in the abiraterone group were classified as low-volume, although the PEACE-1 trial did not demonstrate a survival benefit (HR 0.83) in the low-volume population. Stratifying patients according to selected ARPI, 79.4% of patients were treated with abiraterone, what we expected as data of the ARASENS trial was published almost 7 months after PEACE-1. Interestingly, we observed that since the EMA approval of darolutamide in triplet therapy, 71% of patients were treated with darolutamide and 29% with abiraterone, respectively, suggesting that darolutamide is the preferred ARPI. Concerning the use of abiraterone generics, only half of the patients switched on a generic agent since its introduction at the end of 2022.

Usually, elderly patients are often underrepresented in phase III trials, therefore not reflecting the use of the study medication in the effective patient population. Indeed, the median patient age of our real-world population was 68 years (range 49-93 years) and thus comparable with the 2 RCT (median 67 years), confirming that triplet therapy is preferably offered to younger patients. However, our collective comprised even 4 patients ≥80 years. Of note neither PEACE-1 nor ARASENS performed subgroup analysis stratifying treatment efficacy according to age. Thus we recently conducted a network meta-analysis descrying that triplet therapy (ADT plus darolutamide plus docetaxel) may be the most preferred combination even for mHSPC patients ≥65 years in computational models (14).

Evaluating efficacy of triplet therapy, we observed impressive PSA and imaging-based response rates with no significant differences among cohorts. As expected, synchronous metastatic patients have worse survival rates compared to those with metachronous disease. Furthermore, we aimed to depict which and to what extent deviations from the pivotal studies in terms of treatment regimens are performed in the real-world setting. While the administered drug doses were comparable to both phase III trials, we observed protocol deviations concerning time intervals from chemotherapy to ARPI and vice versa, ranging from the recommended simultaneous start towards a delay of >28 days with chemotherapy as the last introduced agent.

Of importance, the time delay of the simultaneous onset of chemotherapy and ARPI was significantly associated with higher rates of disease progress (p=0.015). Assessing the sequence, starting ARPI before chemotherapy was associated with significant higher progression probability (p=0.023), while vice versa had no influence on treatment response (p=0.256). Yet this finding is in contrast to a post HOC analysis of the TITAN trial, where 10.7% of high-volume mHSPC patients were treated with up to 6 cycles docetaxel within two months prior the randomization to ADT plus apalutamide vs. control demonstrating that prior use of docetaxel did not further improve survival or PSA response (15).

Remarkably among the 21.4% of patients with metachronous disease, 81.3% were treated with abiraterone although in contrast to ARASENS, in PEACE-1 exclusively synchronous mHSPC patients were enrolled.

Of importance, approximately 25% of bone metastatic patients were treated with denosumab 120 mg/4 weekly. This is quite surprising as there is no guideline recommendation for the use of skeletal related events prevention in mHSPC as the results of STAMPEDE did not show a benefit of zolendronic acid in this clinical stage and data regarding denosumab in mHSPC is still missing (16). In line, a recent retrospective trial reported that 23.6% of mHSPC patients are treated with bone-modifying agents (BMA), with a predominate use of denosumab, and oncologists being the primary treating physicians prescribing osteoprotective agents (17). In contrast, a recent German questionnaire-based study involving 3692 patients reported that only 1/5 patients with non-metastatic HSPC receiving ADT are treated with BMA (18). Unlike in both RCTs, only 50% of patients sustained prophylactic gCSF 24-hours after application of chemotherapy, resulting in neutropenia in 10/96 patients encompassing six patients with febrile neutropenia.

Summarizing, even though two patients died of multi-organ failure during therapy, we claim that triplet therapy is satisfiable tolerable with manageable side effects. Furthermore, most AEs are related to docetaxel treatment. Even if the darolutamide group was very small, we observed only two severe side effects, the multi-organ failure and neutropenia. This leads us to the suggestion that darolutamide might be better tolerable than abiraterone in triplet therapy, which has to be further evaluated in a larger collective.

In line with study protocols, all our patients continued ARPI therapy even when exceptional response rates (imaging and PSA based) were achieved. However, there is an ongoing discussion if, in patients with exceptional response rates, ARPI treatment can be reduced or even stopped until relapse. In addition, more and more trials in different settings try to waive classical ADT when ARPI is administered. Thus, we emphasize the importance of clinical trials dealing with these issues.

Both PEACE-1 and ARASENS used conventional imaging methods (CT, bone scan, magnetic resonance imaging) for determination of metastatic load. Generally, the introduction of PSMA PET-CT had a substantial impact on the management of PCa patients with a stage migration phenomenon towards an aggressive phenotype. Current guidelines are not consistent in terms of PSMA PET-CT in the primary staging: While the NCCN (Ver 1.2023) or ESMO guidelines recommend the use in high risk PCa defined as ≥pT2c or Gleason Score 9-10 or PSA >20ng/ml, recommendation of the EAU highlight that although PSMA PET-CT increases the sensitivity, there is lack of outcome data of subsequent treatments based on this imaging method and treatment decisions should not be based on PSMA PET-CT (8,19,20).

Regarding our population, 51% of patients underwent a PSMA PET-CT in the primary staging. Of note, 6 patients were staged by both conventional imaging and PSMA PET. This finding reflects that in countries, like Austria where PSMA PET-CT is an available, easily accessible and financed by the health system, it is the preferred staging method.

Finally, we have to state some limitations: Ⅰ) the study is retrospective and based on 97 Caucasian males from one single European country; Ⅱ) in our cohort abiraterone was used predominantly as selected ARPI possibly causing a statistical bias; Ⅲ) follow-up is short and does not include the same period in all patients with information on castration resistance and OS available only in a very small cohort. Thus, re-analyzing the same cohort in 2-3 years might be highly informative.

## 5. Conclusions

In the present study we assessed the use, performance and tolerability of treatment intensification consisting of ADT plus ARPI plus docetaxel in mHSPC in the real-world setting. Our results support the use of ARPI (abiraterone or darolutamide) in combination with ADT and docetaxel in patients with mHSPC and highlight the importance of the simultaneous start of substances.

## 6. Tables

**Supplementary Table 1:** Clinical parameters at diagnosis stratified according to selected ARPI treatment.

| Parameter | Abiraterone | Darolutamide | p-value |
| --- | --- | --- | --- |
| **Age: Median (SD)** |  |  |  |
| Total |  |  | 0.378 |
| - >65 years | 67 (7) | 70 (10) |  |
| - <65 years | 43 (33) | 11 (6) | 0.597 |
| **PSA at diagnosis (ng/ml) Median (SD)** | 56 (4074) | 60.3 (523) | 0.812 |
| **ISUP grade N (%)** |  |  |  |
| - 1-3 | 10 (13) | 3 (17.6) | 0.136 |
| - 4-5 | 67 (87) | 14 (82.4) |  |
| **Volume of disease *N* (%)** |  |  |  |
| - High | 65 (84.4) | 13 (76.5) | 0.482 |
| - Low | 12 (15.6) | 4 (23.5) |  |
| **Metastatic stage *N* (%)** |  |  |  |
| - Synchronous | 66 (85.7) | 14 (82.4) | 0.713 |
| - Metachronous | 11(14.3) | 3 (17.6) |  |
| **Metastasis location *N* (%)** |  |  |  |
| - Singular | 18 (23.4) | 7 (41.2) | 0.06 |
| - Multiple | 59 (76.6) | 10 (58.8) |  |
| **Imaging response *N* (%)** |  |  |  |
| - Stable disease/ Response | 51 (88) | 6 (75) | 0.06 |
| - Progressive disease/ Mixed response | 7 (12) | 2 (25) |  |

## Data Availability

All data produced in the present study are available upon reasonable request to the authors

## Acknowledgments

None

